# Wastewater monitoring outperforms case numbers as a tool to track COVID-19 incidence dynamics when test positivity rates are high

**DOI:** 10.1101/2021.03.25.21254344

**Authors:** Xavier Fernandez-Cassi, Andreas Scheidegger, Carola Bänziger, Federica Cariti, Alex Tuñas Corzon, Pravin Ganesanandamoorthy, Joseph C. Lemaitre, Christoph Ort, Timothy R. Julian, Tamar Kohn

## Abstract

Wastewater-based epidemiology (WBE) has been shown to coincide with, or anticipate, confirmed COVID-19 case numbers. During periods with high test positivity rates, however, case numbers may be underreported, whereas wastewater does not suffer from this limitation. Here we investigated how the dynamics of new COVID-19 infections estimated based on wastewater monitoring or confirmed cases compare to true COVID-19 incidence dynamics. We focused on the first pandemic wave in Switzerland (February to April, 2020), when test positivity ranged up to 26%. SARS-CoV-2 RNA loads were determined 2-4 times per week in three Swiss wastewater treatment plants (Lugano, Lausanne and Zurich). Wastewater and case data were combined with a shedding load distribution and an infection-to-case confirmation delay distribution, respectively, to estimate incidence dynamics. Finally, the estimates were compared to reference incidence dynamics determined by a validated compartmental model. Incidence dynamics estimated based on wastewater data were found to better track the timing and shape of the reference infection peak compared to estimates based on confirmed cases. In contrast, case confirmations provided a better estimate of the subsequent decline in infections. Under a regime of high-test positivity rates, WBE thus provides critical information that is complementary to clinical data to monitor the pandemic trajectory.

## 1. Introduction

Wastewater-based epidemiology (WBE), a form of environmental surveillance of infectious diseases, has long been suggested as a sensitive tool to monitor pathogen circulation in a population^1–3^. Many pathogens, both enteric and otherwise, are excreted from infected individuals into the sewage system via feces, saliva or other bodily fluids^1^. The principle underlying WBE is that the pathogen concentrations or loads in sewage are proportional to the number of infected individuals among the population contributing to the sewage, and can thus inform on the presence and trajectory of a disease outbreak. For example, norovirus concentrations in sewage were found to closely track the dynamics gastroenteritis cases over several years in Japan^4^. WBE can inform not only on the presence and dynamics of a pathogen but may also capture the emergence of new strains or variants before they become widespread in a population^4,5^.

WBE has received renewed attention during the COVID-19 pandemic, when it was recognized that SARS-CoV-2 RNA is excreted in feces ^6^ and can be detected in wastewater^7–9^ and sludge^10,11^. Several studies have shown that the dynamics of SARS-CoV-2 RNA in raw wastewater or sludge coincide with, or even anticipate, the dynamics of confirmed cases^7,10,11^. In addition, WBE was able to capture the introduction and spread of SARS-CoV-2 variants of concern^12^, and identify mutations that were not captured in clinical samples^13^. WBE may thus serve as a useful tool to support COVID-19 monitoring, and WBE data have already been integrated into multiple national or local COVID-19 dashboards^14–17^.

While WBE will never replace case reporting, it can be used to strengthen the understanding of infectious disease dynamics as it holds important benefits over clinical tests. Specifically, WBE captures both symptomatic and asymptomatic virus shedders; WBE data are not affected by testing capacity, strategy or compliance; and WBE allows the monitoring of a large population with few samples. The advantages of WBE over clinical testing are particularly important when test capacity is exceeded and hence may be insufficient to accurately capture case numbers. According to the WHO, the test positivity rate should remain < 5% to confidently track disease dynamics^18^. Under regimes with a positivity rate > 5%, WBE may thus better reflect true disease dynamics than clinical case numbers.

In this study we evaluated the use of wastewater monitoring as a tool to track COVID-19 dynamics. We hypothesized that under high test positivity rates, wastewater provides an improved estimate of the dynamics of new infections (incidence dynamics) compared to confirmed case numbers. We focused on the first wave of the COVID-19 pandemic in Switzerland, which lasted from late February to April 2020. The test positivity rate during this period ranged up to 26 % (Figure S1). Wastewater was monitored 2-4 times per week in two locations (Lugano, Lausanne) that were strongly affected, and one location (Zurich) that experienced a milder wave. We did not directly evaluate SARS-CoV-2 RNA loads measured in wastewater against the number of confirmed cases. Instead, we use these metrics to estimate the incidence dynamics over time. This allowed us to compare both the wastewater- and the case number-derived incidence dynamics to reference incidence dynamics determined retrospectively by a compartmental (Susceptible-Exposed-Infected-Recovered; SEIR) model and consistent with seroprevalence studies conducted in the region^19^.

## 2. Materials and Methods

### 2.1. Experimental approach

We determined the concentration and daily loads of SARS-CoV-2 RNA in longitudinal samples of raw wastewater collected from three Swiss wastewater treatment plants (WWTPs). In each sample, we analyzed two SARS-CoV-2 gene targets (N1 and N2). In addition, we determined virus recovery by means of an externally added viral surrogate of SARS-CoV-2. Finally, we monitored the fecal strength in each sample via the analysis of pepper mild mottle virus (PMMoV), a plant virus that occurs in wastewater at high and constant concentrations^20,21^.

### 2.2. Sample collection and storage

24-h composite influent samples were collected 2-4 times per week between February 26 and April 30, 2020 from three Swiss WWTPs: Lausanne (STEP de Vidy; population connected: 240’000; 25 samples), Lugano (CDA Bioggio; population connected: 125’000; 31 samples); and Zürich (ARA Werdhölzli: population connected: 450’000; 22 samples). After collection, the wastewater samples were stored at - 20 ºC for up to 5 months.

### 2.3. Preparation of viral surrogate stock solutions

Three enveloped viruses were assessed as external recovery controls, namely Murine Hepatitis Virus (MHV, *Coronaviridae, betacoronavirus*), *Pseudomonas virus* Φ6 (*Cystoviridae, cystovirus*) and Murine Sendai virus (*Paramyxoviridae, respirovirus*). Murine Hepatitis Virus strain MHV-A59 (kindly donated by Volker Thiel, University of Bern) was propagated in DBT cells (kindly donated by Krista Rule Wigginton, University of Michigan) as described elsewhere^22^. Five days post-infection the viral particles were released from infected cells by three cycles of freezing/thawing. Cell supernatants were centrifuged at 3000 x*g* to pellet down cell debris and the supernatant was clarified through a 0.22 µm filter. The resulting stock solution had a concentration of 7.8×10^9^ genome copies (gc)/ml. Bacteriophage Φ6 (DSMZ nº 21518, strain HER 102) was propagated in *P. syringae* (DSMZ nº 21482, strain HER1102) according to the provider’s instructions. After propagation, bacterial cultures were centrifuged at 8000 x*g* for 10 min and cell debris was removed by passing the supernatant through a 0.22 µm filter. The final stock solution had a concentration of 8.0×10^8^ gc/ml. Finally, Sendai virus propagated in embryonated eggs was kindly donated by Dominique Garcin (University of Geneva) and was used without further treatment. These solutions had a concentration of 1.3×10^9^ gc/ml.

### 2.4. Sample concentration and nucleic acid extraction

Samples from Lugano and Lausanne were processed at EPFL, and samples from Zürich were processed at Eawag. Prior to processing, samples were thawed at room temperature. For each sample, two replicate aliquots of 50 ml wastewater were processed. The 50 ml aliquots were spiked with MHV (Lausanne or Lugano) or Sendai virus (Zürich) at a concentration of approximately 1×10^6^ gc/50 ml, and were stirred for 20 minutes to ensure the homogenization of the sample. Then they were pre-filtered using 2 µm glass fiber pre-filters (Merck, cat nº AP2007500) placed on the top of 0.22 µm SteriCup filters (Merck, cat nº SCGVU02RE). After filtration, the filter units were rinsed with 10 ml of ultrapure water to ensure that no wastewater was retained in the dead volume. The filtrates (approximately 60 ml) were transferred to a centrifugal filter unit with a size cut-off of 100 kDa (Centricon Plus-70; Millipore cat nº UFC701008), and were centrifuged for 30 min at 3000x*g*. To collect the concentrate, the centrifugal filter was inverted and centrifuged for 3 min at 1000x*g*. The resulting viral concentrate volume ranged from 180 to 300 µL.

Viral concentrates were extracted in their entirety using the Qiagen RNA Viral Mini Kit (Qiagen cat nº 22906, Valencia, CA, USA) following the manufacturer’s protocol for higher volumes. Nucleic acids were eluted using 80 μl of AVE buffer. For each processed batch of samples, a negative extraction control using water was included. The extracted nucleic acids were passed through a Zymo OneStep PCR Inhibitor Removal column (Zymo Research, cat nº D6030) to remove PCR inhibitors following the protocol provided by the manufacturer.

In addition to the longitudinal samples, seven wastewater samples were collected in Lausanne to test the recovery of different SARS-CoV-2 surrogates. These samples were spiked with MHV, Sendai virus and Φ6 at a concentration of approximately 10^6^ gc/50 ml each. Samples were then concentrated and nucleic acids were extracted as described above.

### 2.5. Quantification of SARS-CoV-2 N1 and N2 genes, viral surrogates and PMMoV by RT-qPCR

All RNA extracts of the longitudinal samples were analyzed by RT-qPCR for four viral targets: The N1 and N2 genes of SARS-CoV-2, the surrogate virus and PMMoV. All N1, N2 and MHV analyses, as well as PMMoV analyses for Lugano and Lausanne were performed at EPFL. PMMoV and Sendai virus analyses for Zürich were performed at Eawag. The samples to test surrogate virus recovery were analyzed at EPFL for three viral targets: MHV, Sendai virus and Φ6. To detect the presence of SARS-CoV-2 RNA, the CDC N1 and N2 assays were used^23^. PMMoV and MHV were analyzed by previously reported assays^24–26^. The design for primers and probes for Φ6 were adapted from Gendron et al^27^ according to the suggestion of Heather Bischel (University of California, Davis). For Sendai virus, primers and probes were designed for the purpose of this project. A summary of all primers and probes and the RT-qPCR protocols is given in the Supporting Information (Table S1).

To calibrate the different RT-qPCR assays, standard curves for each viral target were generated using either double-stranded DNA gblocks gene fragments (viral surrogates and PMMoV), or a 2019-nCoV_N positive control plasmid (cat nº 10006625, SARS-CoV-2 N1 and N2). Both gBlocks and plasmids were purchased from Integrated DNA Technologies (Coralville, IA, USA).

RT-qPCR amplifications were performed in 25 µl reactions using RNA UltraSense™ One-Step Quantitative RT-PCR System (Invitrogen, cat nº 11732-927), amended with 4 µl of bovine serum albumin (2 mg/ml) on a Mic qPCR Cycler (Bio Molecular Systems). In each RT-qPCR reaction, 5 µl of RNA extract or calibration standard were used. For PMMoV, the RNA extract was diluted 1:10 prior to RT-qPCR analysis. All RT-qPCR runs included no-template controls and negative extraction controls to monitor for contamination during the extraction and amplification process. The preparation of PCR mastermix and standards, as well as sample loading were performed in separate locations to avoid contamination. Cq determination was performed using the micPCR software (v2; Bio Molecular Systems).

RT-qPCR limits of detection (LOD) were determined as the lowest concentration (N1, N2 and MHV) or the lowest standard (PMMoV, Sendai, Φ6) with a 95% or greater detection probability. The limit of detection (LOD) for each gene target were determined from pooled standard curves (n ≥ 3) in R using the Generic qPCR Limit of Detection (LOD) / Limit of Quantification (LOQ) calculator^28^. Samples with a measurable RT-qPCR signal < LOD were assigned the concentration of the LOD of the respective assay. Samples which yielded no detectable RT-qCR signal were set to the theoretical minimal LOD (3 gc/reaction)^29^.

### 2.6. RT-qPCR inhibition

To check for inhibition during RT-qPCR reactions, 4 µl of each zymo-treated RNA extract were amended with 1 µl of a synthetic SARS-CoV-2 RNA reference material (EURM-019, Joint Research Center) at a concentration of approximately 10^5^ gc/µL. RNA extracts were analyzed for the SARS-CoV-2 N1 target by the RT-qPCR protocol described above, and the resulting Cq values were compared between samples. Samples were considered inhibited when Cq was > 1.5 cycles beyond the average Cq measured at a given site. All inhibition tests were conducted at EPFL.

### 2.7. Recovery

Recovery was calculated as the ratio of surrogate virus recovered after sample processing and the virus originally spiked into 50 ml of unfiltered wastewater (equation 1):

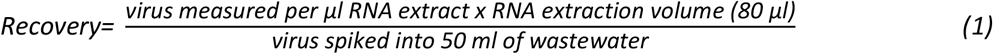

### 2.8. Determination of RNA loads

Genome copies (gc) per reaction were converted to units of load (gc/day) by determining the gc concentration per liter of wastewater and multiplication by the wastewater flow rate of corresponding day according to equation 2:

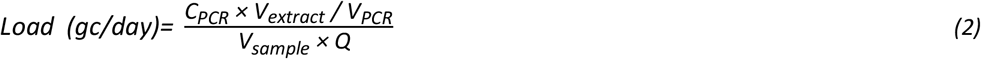

Where C_PCR_ is the template concentration (gc/reaction) determined by RT-qPCR, V_extract_ is the total volume of RNA extract (80 μL), V_PCR_ is the volume of extract analyzed by RT-qPCR (5 μL), V_sample_ is the volume of the wastewater sample (0.05 L), and Q is the wastewater flow rate on a given sampling day measured and provided by the WWTPs included in this study (L/day).

### 2.9. Storage test

To determine if storage at -20 °C had a detrimental effect on SARS-CoV-2 RNA concentrations in wastewater, a control experiment was conducted. A batch of wastewater influent from Lausanne was collected and stored for a month in four different conditions: 1) unprocessed wastewater at 4 °C; 2) unprocessed wastewater at -20 °C; 3) concentrated wastewater (after filtration and ultracentrifugation) at -20 °C; and 4) zymo-treated RNA extract at -20 °C. All tests were conducted in duplicate. After one month, all samples were fully processed and immediately analyzed for the N1 gene target by RT-qPCR as described above.

### 2.10. Epidemiological data

Confirmed case numbers for each WWTP catchment were kindly provided by the Swiss Federal Office of Public Health.

### 2.11. Incidence estimates

Reference infection numbers were determined by an SEIR model described previously ^19^. This model is based on cantonal hospitalization data, intensive care unit visits and deaths, but not case numbers. This is to avoid any influence from changes in test strategies and test capacity over the time period considered. The modeled incidence includes both symptomatic and asymptomatic new infections. The model was validated against a seroprevalence study conducted in the region, and we therefore consider it herein as the reference incidence.

Incidence was additionally estimated based on longitudinal data of SARS-CoV-2 loads in wastewater and based on confirmed cases. Both these metrics measured at time *t* reflect an aggregate of infections that occurred over a time span preceding the measurement. A deconvolution of these aggregated quantities allows for the reconstruction of daily infections. This requires an assumption of the extent by which previous infections influence the aggregated quantities. This assumption is expressed by time delay-dependent weight *ω(τ)*, where *τ* is the time (days) since infection. The measured aggregated quantity *A* at time *t* can be approximated by a weighted sum of all infections *I* that occurred up to time *t*:

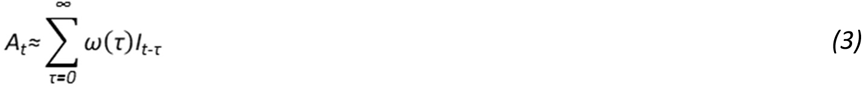

The infections *I* over a time range of interest can be estimated via non-negative least squares regression. As fast fluctuations in the number of daily infections seems unreasonable, an additional constraint was added to enforce smoothness comparable to the infection numbers of the SEIR model (see Supporting Information).

To obtain the wastewater-derived incidence, the weights of the deconvolution, *ω(τ)*, are given by the shedding load profile (SLP) that describes the average amount of virus shed by a patient *τ* days after infection. The SLP can be decomposed into the relative shedding load probability distribution (SLD) and the absolute viral load shed during the course of the disease (L) (equation 4):

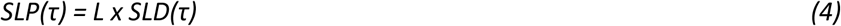

The SLD was constructed by combining the gastrointestinal viral load as a function of time after symptom onset with the time between infection and symptoms. Virus shedding was modelled based on data reviewed by Benefield et al.^30^, and could be well described by a gamma distribution with a mean of 6.73 days and a standard deviation (sd) of 6.98 days. The time between infection and symptoms was also modeled by a gamma distribution based on Linton et al.^31^ (mean = 5.3 days, sd = 3.2 days). The convolution of these two distributions was used as the SLD (shown in Figure 1a) with a mean = 11.73 days and an sd = 7.68 days.

**Figure 1.**
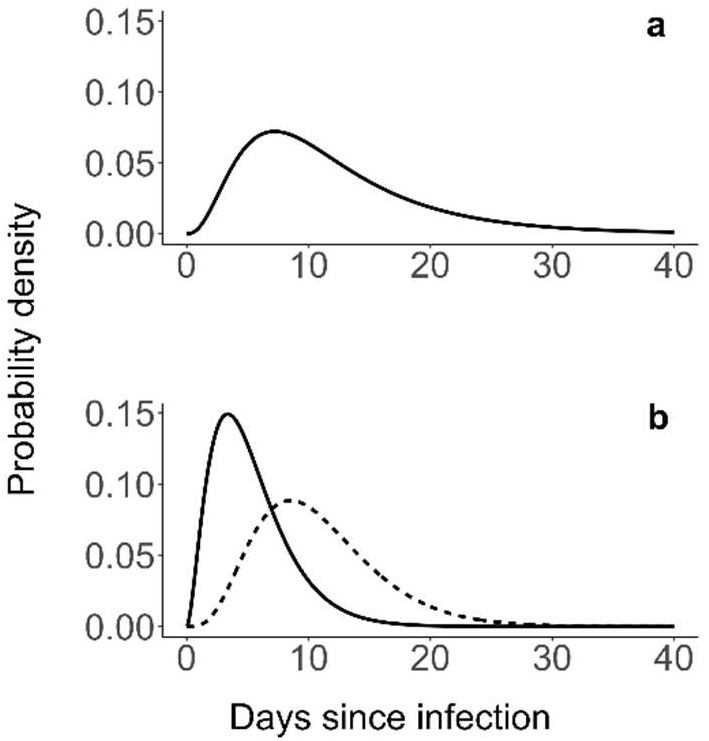
a) Shedding load distribution to describe the relative distribution of the SARS shedding load, based on the delay distribution from infection to symptom onset by Linton et al.^31^, combined with the gastrointestinal viral load dynamics according to Benefield et al.^30^ b) Delay distribution from infection to symptom onset according to Linton et al.^31^ (solid line), and combined with an additional delay from symptom onset to case confirmation based on Bi et al.^34^ (dashed line).

If L is known with small uncertainty, the absolute number of infections can be estimated. However, although the different SLDs have a comparable shape across literature, the loads L are highly variable ^32,33^. Therefore, we applied the SLD, which still yields an estimate that is proportional to that obtained by using the correct but unknown SLP.

For the case number-derived incidence, *(τ)* was defined by the distribution combining the delays from infection to symptom onset (gamma mean= 5.3 days; sd = 3.2 days), and from symptom onset to case confirmation (gamma mean= 5.5 days, sd = 3.8 days^34^). The resulting delay distribution from infection to case confirmation is visualized in Figure 1b.

### 2.12. Data analysis

All statistical analyses were performed in R^35^. The non-negative least-square regression for the incidence estimations was implemented with the package ‘CVXR’ ^36^ and delay distributions were computed with the R package ‘distr’^37^.

### 2.13. Data availability

Data will be made available on an institutional repository upon acceptance of the manuscript.

## 3. Results and Discussion

### 3.1. Method performance

#### 3.1.1. PCR efficiency and LOD

PCR efficiencies for all targets ranged from 94-111% (Table S2). The R^2^ of the pooled standard curves were ≥ 0.95. No amplification signal was measured in the non-template and negative extraction controls confirming the absence of contamination during sample processing. The LOD corresponded to 4.2 gc/ml wastewater and 2.6 gc/ml wastewater for the SARS-CoV-2 N1 and N2 genes, respectively. The LODs reflect the difficulty of producing accurate calibration curves for SARS-CoV-2 in the low template range based on plasmid standards. This limitation, which was also reported by others ^38,39^ highlights the need for improved qPCR standards and more sensitive RT-qPCR assays to minimize variability and false negative results in SARS-CoV-2 RNA quantification. The LODs of the other targets are listed in Table S2.

#### 3.1.2. PCR Inhibition

Spiking RNA extracts with synthetic SARS-CoV-2 RNA reference material revealed minimal PCR inhibition on most samples. Specifically, with the exception of three samples from Lugano, all spiked RNA extracts exhibited N1 Cq values that fell within or only minimally beyond 1.5 cycles of the median Cq of a given WWTP (Figure S2).

#### 3.1.3. Reproducibility

We compared quantifiable N1 concentrations determined in biological as well as in technical replicate samples (Figure S3). A good correlation (*r*=0.89) was obtained among biological replicates, indicating a high reproducibility of the overall processing pipeline. A good reproducibility was also found for technical replicates (*r*=0.78).

#### 3.1.4. Recovery

Three enveloped viruses - MHV, Sendai virus and Φ6 - were evaluated as SARS-CoV-2 surrogates to monitor virus recovery in our sample processing pipeline. As a member of the *Coronaviridae* family, MHV is the most similar to SARS-CoV-2 in terms of size (120 nm diameter) and genome structure (single-stranded RNA). Sendai virus has a single-stranded RNA genome, but is slightly larger in diameter than SARS-CoV-2 (150 nm). Besides SARS-CoV-2, this virus may also serve as a surrogate for viruses with pandemic potential in the *Paramyxoviridae* family, such as measles virus. The novel RT-qPCR assay developed herein was able to quantify its concentration down to an LOD of 4.2 gc/ml (Table S2). Finally, Φ6 is the least similar surrogate to SARS-CoV-2. It has a smaller diameter (85 nm) and a different genome structure (double-stranded RNA).

In seven wastewater samples spiked with all three surrogate viruses, MHV and Sendai virus exhibited similar recoveries that mostly ranged from 0.1-1% (Figure S4). This range corresponds well to that reported by other groups using a similar processing pipeline ^40^. If determined using Φ6, recoveries were 10-to 100-fold higher and more constant across samples. This confirms previous reports that recoveries depend strongly on the surrogate virus used^40^. Despite the better recovery of Φ6, we decided to utilize MHV or Sendai virus as recovery controls in this work, due to their higher structural similarity with SARS-CoV-2.

The recoveries in the samples from Lugano and Lausanne were determined using MHV (Figure S5). Recoveries were similar for both sites and mostly fell into the 0.1-1% range, with average values of 0.95% and 0.74% for Lugano and Lausanne, respectively. In the samples from Zurich, Sendai virus was used as the surrogate. Compared to Lugano and Lausanne, the recoveries were significantly lower, with an average of 0.17% (one-way ANOVA, F=6.82, p < 0.002) (Figure S5). The lower recovery is unlikely to be a result of the use of Sendai virus, since MHV and Sendai virus yielded similar recoveries if assessed in the same sample (Figure S4). Instead, the lower recoveries in the Zurich samples may reflect the higher solids content in this WWTP. Enveloped viruses partition to wastewater solids^41^, and hence a higher solids content leads to a reduced recovery of the surrogate virus from the liquid wastewater fraction.

#### 3.1.5. Fecal load

The daily load of PMMoV was used as an indicator of the fecal load entering the WWTP. On average, the PMMoV loads corresponded to 8.9×10^15^ gc/day (Lugano), 3.1×10^16^ gc/day (Lausanne) and 1.8×10^16^ gc/day (Zurich) (Figure S6). In all but five samples the PMMoV load fell within the range of 5×10^15^ to 5×10^16^ gc/day. The narrow range of the measured PMMoV loads further confirms the consistency of our virus concentration and extraction process.

#### 3.1.6. Effect of storage conditions on RNA stability

As shown in Figure 2, different storage procedures exert significantly different effects on SARS-CoV-2 RNA stability (one-way ANOVA, F=12.8, p < 0.001). Storing raw wastewater at 4 °C or -20 °C for a month resulted in lower concentrations of SARS-CoV-2 RNA compared to samples stored as concentrates or RNA extracts at -20 °C (Tukey-Kramer, p < 0.02). The storage protocol used herein (raw wastewater at -20 °C) had to be implemented before storage tests could be completed, and likely led to significant RNA decay. In future studies, wastewater samples should immediately be concentrated or extracted prior to storage.

**Figure 2.**
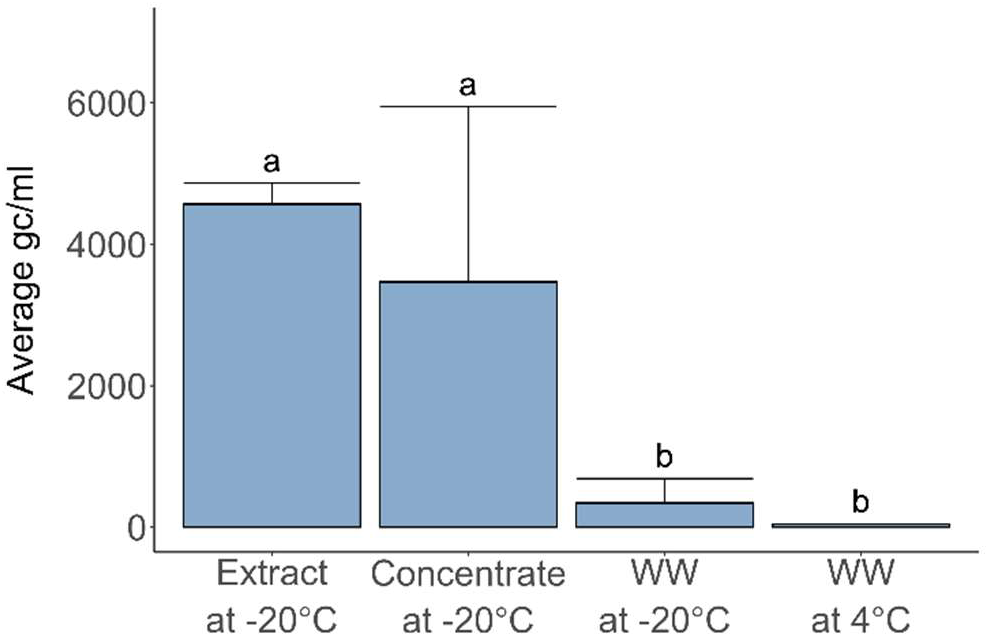
Effect of sample storage over one month under different conditions on SARS-CoV-2 RNA concentrations (N1 gene, gc/ml wastewater). Error bars represent standard deviations of replicate samples. Samples stored as non-processed raw wastewater (WW) at 4 °C or -20 °C exhibited lower concentrations compared to samples stored at -20 °C as concentrate (post ultrafiltration) or RNA extracts. Indices a and b denote experimental conditions yielding statistically different sample means.

### 3.2. Longitudinal trends SARS-CoV-2 RNA loads and confirmed cases

Concentrations of SARS-CoV-2 N1 and N2 genes in longitudinal samples are shown in Figure S7. The earliest detection of the N1 gene occurred in wastewater from Lugano on February 28 2020, four days after the first case was observed in Switzerland. This confirms earlier reports that wastewater can serve as a sensitive indicator for virus circulation, even during periods of low disease prevalence ^7,8,42,43^.

N1 and N2 concentrations exhibited similar temporal trends, though N1 concentrations were on average 3-fold higher (Figure S7). Higher concentrations of the N1 gene were also reported by others ^38,44^, though some studies have reported the N2 gene to yield higher results^7,45^. Given the superior quantification by N1 in this work, only this gene target was considered for all subsequent analyses.

We did not normalize N1 concentrations by fecal strength (PMMoV concentration) as suggested elsewhere^44,46^, because PMMoV concentrations were similar in all samples and were not correlated with N1 concentrations (*r*=0.02-0.04) (Figure S8). We also did not correct N1 concentrations for recovery, because there was high inter-sample variation across the three surrogates tested, and it is uncertain which - if any - externally added surrogate accurately mimics the fate of SARS-CoV-2 during sample processing^11,39^. Recovery values were strictly used for data quality control. Specifically, samples were excluded if two criteria were simultaneously met: the recovery of a sample was > 3x the median recovery for the site under consideration; and the concentrations measured by N1 and N2 differed by more than a factor 5. This led to the exclusion of one biological replicate on three sampling days in Lugano (March 18-19 and April 5), and both replicates for a single day in Lausanne (March 28).

Among the three WWTPs studied, Lausanne had the highest number of confirmed cases in its catchment (Figure 3). Case numbers were similar in the catchments of the Lugano and Zurich WWTPs, even though Zurich’s catchment encompasses approximately 3.6-fold more inhabitants than Lugano’s, and 1.9-fold more than Lausanne’s. Consequently, the N1 concentrations in the Zurich WWTP were expected to be lower compared to Lugano and Lausanne, as confirmed by our measurements (Figure S7). To enable a direct comparison among WWTPs, we converted N1 concentrations into units of daily N1 load (equation 2). This unit accounts for differences in catchment size (via the daily wastewater flow rate), and also incorporates daily variability in the wastewater flow of a given WWTP.

**Figure 3.**
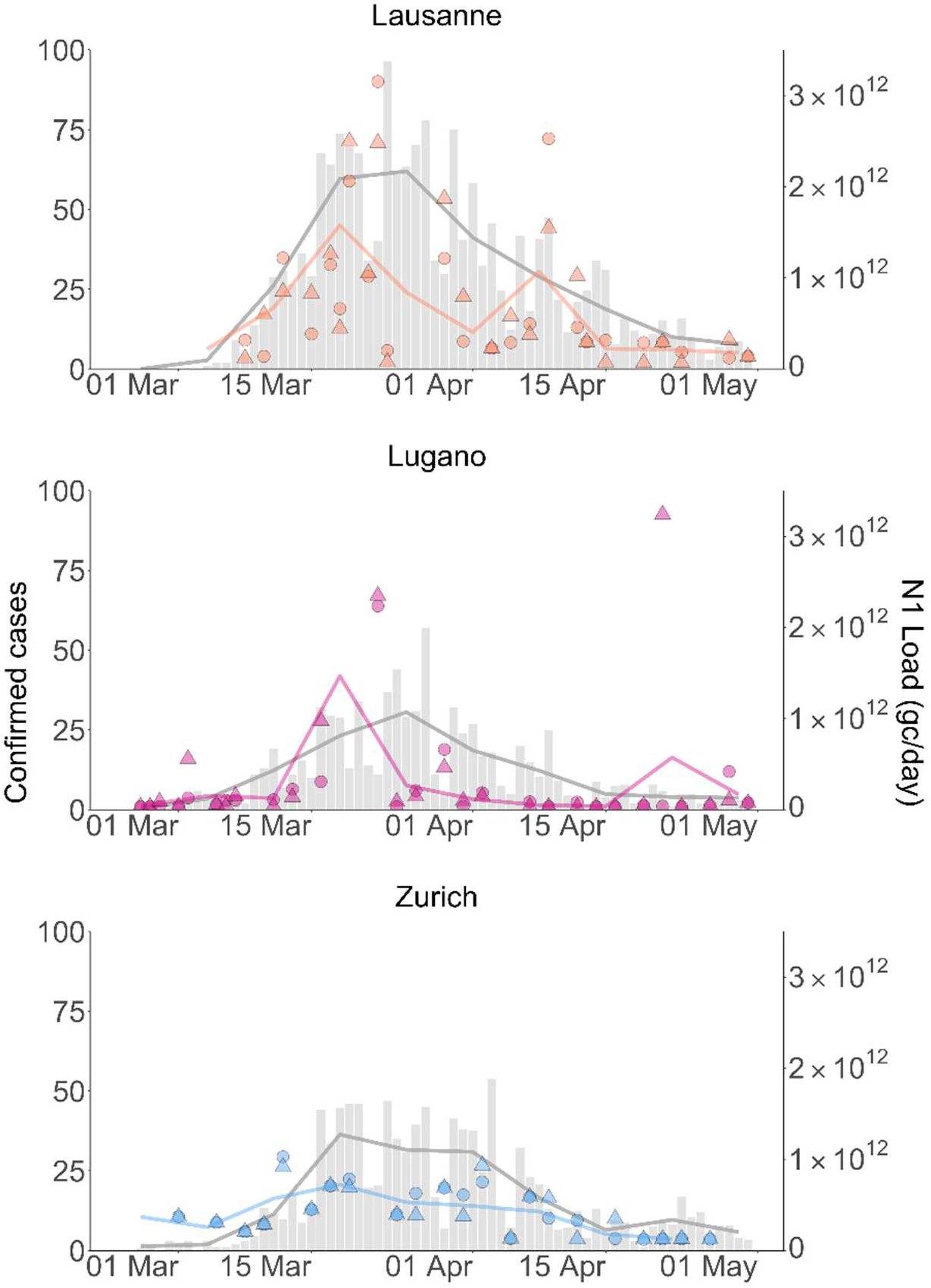
SARS-CoV-2 RNA (N1) loads and confirmed cases for the Lausanne, Lugano and Zurich WWTP catchments from February 26 until April 30, 2020. Data points represent wastewater data (average of technical replicates). Circles and triangles indicate biological replicates. Grey bars show confirmed cases. Lines connect weekly (Monday-Sunday) averages of SARS-CoV-2 RNA loads or confirmed cases.

Similar to data from other studies^10,11,38^, there was considerable day-to-day variability in both the N1 loads and the number of confirmed cases (Figure 3). The variability in confirmed cases is increased by the fact that Switzerland reduces testing and reporting on weekends. To facilitate the visualization of pandemic trends in wastewater and case data, we therefore calculated weekly averages (Monday - Sunday) for each data set. The corresponding results are shown as solid lines in Figure 3. As is evident, both data sets feature a prominent peak in late March. However, the wastewater peak shape is narrow, whereas the number of confirmed cases remained high for 2-3 weeks.

Despite the similarity in confirmed case numbers in the catchment, measured SARS-CoV-2 RNA loads were higher in the Lugano WWTP than in Zurich WWTP. There are a number of potential methodological explanations for this, including lower virus recovery in Zurich (Figure S5), lower precision in quantifying low SARS-CoV-2 RNA copy numbers (Figure S7), and our sample storage protocol, which in retrospect was found to be non ideal (Figure 2). The Zurich WWTP is located in the area of lowest disease prevalence and thus had the lowest starting concentrations of SARS-CoV-2 RNA among the WWTPs sampled. Further decay during storage of the Zurich samples may have lowered the concentrations below the LOD in all but the samples taken during the peak of the first wave.

### 3.3. Comparison of incidence dynamics from wastewater data, case numbers and SEIR models

To assess the ability to track disease dynamics with SARS-CoV-2 loads in wastewater and confirmed cases, both data sets were used to estimate disease incidence over time (by deconvoluting the signals, see Materials and Methods). The resulting trends were compared to the reference incidence determined by an SEIR model ^19^. While the SEIR model reports absolute infection numbers, this determination is currently not feasible for wastewater- or case number-derived estimates. For wastewater, estimating absolute infection numbers would require a better understanding of the magnitude of the shedding load L (equation 4), the decay kinetics of SARS-CoV-2 RNA in the sewer system, and the true recovery of SARS-CoV-2 in our sample processing pipeline. These parameters are currently not available, but may become better known in the future. For case numbers, the ratio of confirmed to total cases would have to be known, yet this parameter is associated with considerable uncertainty and likely variability during the first wave of the pandemic. We therefore only compared the incidence dynamics, but not the absolute incidence per day.

As shown in Figure 4, both wastewater- and case number-derived incidence exhibited a pronounced peak in mid-March. In Lausanne, the wastewater-derived incidence exhibited the highest number of infections from March 13-15, which matches the peak of infections determined by the SEIR model. If estimated based on confirmed cases, the highest number of new infections occurred from March 9-11. Considering the delay distributions from infection to case confirmation, this time range mainly reflects cases observed from March 17-24, coinciding with Swiss-wide positivity rates > 10% (Figure S1). The premature timing of the peak may indicate that case numbers were truncated when testing capacity was exceeded and positivity rates were high.

**Figure 4.**
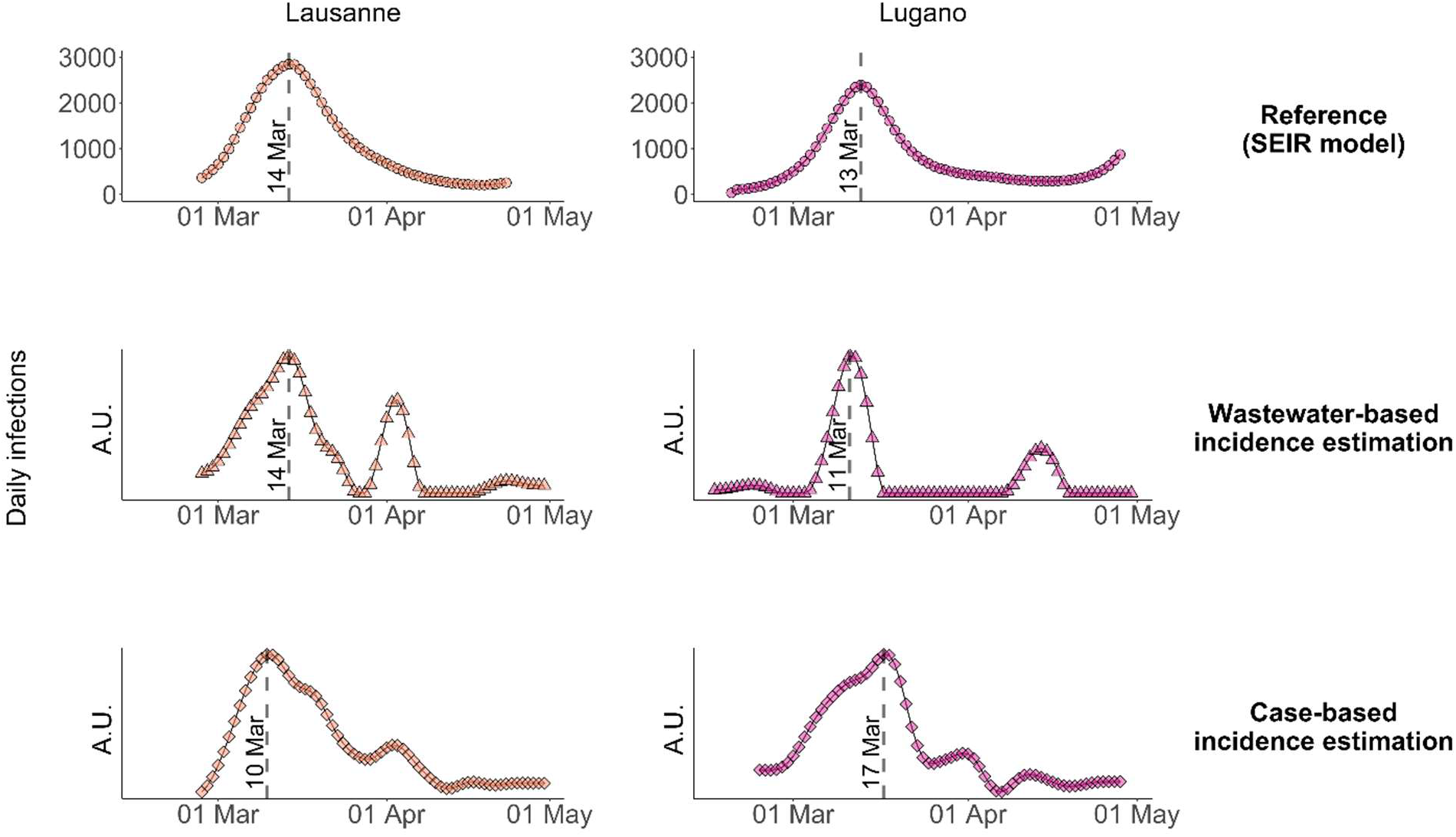
Comparison of COVID19 incidence dynamics estimated by the SEIR model, determined based on SARS-CoV-2 RNA loads in wastewater and based on confirmed case numbers. Incidence dynamics were determined by deconvolution of the wastewater loads and case numbers shown in Figure 3. The Zurich WWTP was not included in this analysis, because most wastewater samples yielded non-detectable SARS-CoV-2 RNA concentrations. A.U. = arbitrary units.

In Lugano, wastewater-based incidence estimates yielded the highest infection numbers from March 10-12. This time frame partly overlapped with the SEIR-modelled infection peak, which occurred from March 12-14. In contrast, the incidence peak determined from confirmed cases appeared later (March 17-19) and differed in shape compared to the other incidence estimates. This is another indication testing capacity during this period was insufficient to capture the full extent of the rise in cases during the height of the first wave.

In both locations, the decline in new infections was better captured by case number-than by wastewater-based incidence estimates. In Lausanne, the case number-based incidence exhibited a slow decay in new infections from mid-March to late April, similar to the reference incidence dynamics. In contrast, the decay in the wastewater-based incidence was faster. In Lugano the case number-derived incidence was also able to capture the tail end of the wave, whereas new infections based on wastewater data rapidly dropped to the baseline.

Finally, the wastewater-based incidence dynamics in both locations exhibited a second, smaller peak in April, which was driven by few high load measurements in each location. In Lausanne this feature also appeared in the corresponding case number-derived incidence dynamics and thus reflects a local spike in infections. In contrast, the origin of the second peak in Lugano is not evident. It may stem from one or more shedders that are not permanent inhabitants of the Lugano WWTP catchment and were therefore not included in the catchment-specific case numbers (e.g., commuters or external patients hospitalized within the catchment). The April peaks were not apparent in the SEIR model, which may be explained by differences in the type of input data used to determine incidence dynamics. Whereas wastewater loads and case numbers were catchment-specific, the SEIR model was based on data for the entire canton. Local spikes in case numbers would thus appear attenuated in the reference incidence.

## 4. Conclusions

Our findings demonstrate that both confirmed case numbers and wastewater analysis are useful and independent metrics to estimate COVID-19 incidence dynamics. Wastewater outperformed case numbers with respect to the timing and shape of the peak incidence, whereas confirmed case numbers were a better indicator for incidence decline. In combination, the two metrics yielded complementary information on incidence dynamics that correspond well to the reference dynamics determined by compartmental models.

It is important to consider that all three approaches rely on a number of assumptions, all of which are associated with a degree of uncertainty. For example, the SEIR model is based exclusively on data pertaining to severe COVID-19 cases (hospitalizations, deaths), and may thus miss events among age classes that have a low severity rate but normal virus shedding. Wastewater-derived incidence dynamics suffer from uncertainties in the accuracy of the SLD. And cases-number derived estimates rely on the delay distribution between infection and case confirmation, which may vary with time and location. While the sources of uncertainties of these assumptions are conceptually understood, they remain difficult to quantify due to the lack of reference data. It is therefore important and encouraging that despite these uncertainties, comparable incidence dynamics were obtained with three independent approaches.

Differences in the incidence dynamics determined by wastewater and confirmed cases may ultimately also be exploited to inform on the duration and degree of clinical undertesting. To do so, however, both incidence estimates need to be further advanced. In future work, wastewater-derived estimates can be enhanced by increasing the wastewater sampling frequency to smooth out measurement outliers, developing more sensitive assays to quantify the viral RNA at low concentrations, better determining SARS-CoV-2 RNA recovery from wastewater, and establishing a representative shedding load profile. Case number-derived incidence estimates can be improved by taking into account variations in the delay distributions from symptom onset to case confirmation. In Switzerland, the mean delay varied from 3 to 8 days during the first wave ^47^, yet herein it was held constant at 5.5 days.

Compared to the compartmental model, which relies on hospitalization and deaths, WBE can determine incidence dynamics with a faster turnaround time (RNA loads can be measured within 24 hours after sampling). Compared to clinical tests, an additional advantage of WBE is that a much lower number of samples is required to determine incidence dynamics with reasonable accuracy. During high positivity rate regimes, WBE can thus yield information on the trajectory of a pandemic that is potentially more precise, more readily available and more economical than information from clinical data. We contend that WBE should be included by epidemiologists and public health agencies as a useful pandemic monitoring tool during periods with high test positivity rates.

## Data Availability

Data will be made available on an institutional repository upon acceptance of the manuscript.

## Acknowledgements

This work was supported by the Swiss National Science Foundation (project 31CA30_196538), Eawag discretionary funds, and an EPFL COVID-19 grant. XFC was a fellow of the European Union’s Horizon 2020 research and innovation programme under the Marie Skłodowska–Curie Grant Agreement No. 754462. We thank the operators of the Lugano, Lausanne and Zurich WWTPs for providing samples, the Swiss Federal Office of Public Health for catchment-specific case numbers, Jana Huisman for valuable input on SLDs, and Marie-Helene Corre, Elyse Stachler and Lea Caduff for lab assistance.

## Appendix A: Supporting Information

Supporting information to this article can be found online at https://doi.org/xxx.

## References

(1) Sinclair, R. G., Choi, C. Y.; Riley, M. R.; Gerba, C. P. Pathogen surveillance through monitoring of sewer systems. Adv Appl Microbiol 2008, 65, 249–269.

(2) Fernandez-Cassi, X.; Timoneda, N.; Martínez-Puchol, S.; Rusiñol, M.; Rodriguez-Manzano, J.; Figuerola, N.; Bofill-Mas, S.; Abril, J. F.; Girones, R. Metagenomics for the study of viruses in urban sewage as a tool for public health surveillance. Sci. Total Environ. 2018, 618, 870–880.

(3) Hovi, T.; Shulman, L. M.; van der Avoort, H.; Deshpande, J.; Roivainen, M.; DE Gourville, E. M. Role of environmental poliovirus surveillance in global polio eradication and beyond. Epidemiol. Infect. 2012, 140, 1–13.

(4) Kazama, S.; Miura, T.; Masago, Y.; Konta, Y.; Tohma, K.; Manaka, T.; Liu, X.; Nakayama, D.; Tanno, T.; Saito, M.; et al. Environmental surveillance of norovirus genogroups I and II for sensitive detection of epidemic variants. Appl. Environ. Microbiol. 2017, 83.

(5) Bisseux, M.; Debroas, D.; Mirand, A.; Archimbaud, C.; Peigue-Lafeuille, H.; Bailly, J.-L.; Henquell, C. Monitoring of enterovirus diversity in wastewater by ultra-deep sequencing: An effective complementary tool for clinical enterovirus surveillance. Water Res. 2020, 169, 115246.

(6) Wu, Y.; Guo, C.; Tang, L.; Hong, Z.; Zhou, J.; Dong, X.; Yin, H.; Xiao, Q.; Tang, Y.; Qu, X.; et al. Prolonged presence of SARS-CoV-2 viral RNA in faecal samples. Lancet Gastroenterol. Hepatol. 2020, 5, 434–435.

(7) Medema, G.; Heijnen, L.; Elsinga, G.; Italiaander, R.; Brouwer, A. Presence of SARS-Coronavirus-2 RNA in Sewage and Correlation with Reported COVID-19 Prevalence in the early stage of the epidemic in The Netherlands. Environ. Sci. Technol. Lett. 2020.

(8) Randazzo, W.; Truchado, P.; Cuevas-Ferrando, E.; Simón, P.; Allende, A.; Sánchez, G. SARS-CoV-2 RNA in wastewater anticipated COVID-19 occurrence in a low prevalence area. Water Res. 2020, 181, 115942.

(9) Ahmed, W.; Angel, N.; Edson, J.; Bibby, K.; Bivins, A.; O’Brien, J. W.; Choi, P. M.; Kitajima, M.; Simpson, S. L.; Li, J.; et al. First confirmed detection of SARS-CoV-2 in untreated wastewater in Australia: A proof of concept for the wastewater surveillance of COVID-19 in the community. Sci. Total Environ. 2020, 728, 138764.

(10) Peccia, J.; Zulli, A.; Brackney, D. E.; Grubaugh, N. D.; Kaplan, E. H.; Casanovas-Massana, A.; Ko, A. I.; Malik, A. A.; Wang, D.; Wang, M.; et al. Measurement of SARS-CoV-2 RNA in wastewater tracks community infection dynamics. Nat. Biotechnol. 2020, 38, 1164–1167.

(11) Graham, K. E.; Loeb, S. K.; Wolfe, M. K.; Catoe, D.; Sinnott-Armstrong, N.; Kim, S.; Yamahara, K. M.; Sassoubre, L. M.; Mendoza Grijalva, L. M., Roldan-Hernandez, L.; et al. SARS-CoV-2 RNA in wastewater settled solids is associated with COVID-19 cases in a large urban sewershed. Environ. Sci. Technol. 2021, 55, 488–498.

(12) Jahn, K.; Dreifuss, D.; Topolsky, I.; Kull, A.; Ganesanandamoorthy, P.; Fernandez-Cassi, X.; Bänziger, C.; Stachler, E.; Fuhrmann, L.; Jablonski, K. P.; et al. Detection of SARS-CoV-2 variants in Switzerland by genomic analysis of wastewater samples. medRxiv 2021.01.08.21249379; doi: https://doi.org/10.1101/2021.01.08.21249379.

(13) Crits-Christoph, A.; Kantor, R. S.; Olm, M. R.; Whitney, O. N.; Al-Shayeb, B.; Lou, Y. C.; Flamholz, A.; Kennedy, L. C.; Greenwald, H.; Hinkle, A.; et al. Genome sequencing of sewage detects regionally prevalent SARS-CoV-2 variants. MBio 2021, 12.

(14) The Cambridge Public Health Department (CPHD). Cambridge COVID-19 Data Center https://cityofcambridge.shinyapps.io/COVID19/#shiny-tab-wastewater (accessed Mar 1, 2021).

(15) Victoria State Government - Health and Human Services. Wastewater monitoring - coronavirus (COVID-19) https://www.dhhs.vic.gov.au/wastewater-monitoring-covid-19 (accessed Mar 1, 2021).

(16) Queensland Government. Wastewater surveillance program results https://www.qld.gov.au/health/conditions/health-alerts/coronavirus-covid-19/current-status/wastewater (accessed Mar 1, 2021).

(17) Rijksoverheid. Early indicators - Virus particles in wastewater https://coronadashboard.government.nl/landelijk/rioolwater (accessed Mar 1, 2021).

(18) World Health Organization. Public health criteria to adjust public health and social measures in the context of COVID-19: annex to considerations in adjusting public health and social measures in the context of COVID-19, 12 May 2020.; WHO/2019-nCoV/Adjusting_PH_measures/Criteria/2020.1; World Health Organization, 2020.

(19) Lemaitre, J. C.; Perez-Saez, J.; Azman, A. S.; Rinaldo, A.; Fellay, J. Assessing the impact of non-pharmaceutical interventions on SARS-CoV-2 transmission in Switzerland. Swiss Med Wkly 2020, 150, w20295.

(20) Kitajima, M.; Iker, B. C.; Pepper, I. L.; Gerba, C. P. Relative abundance and treatment reduction of viruses during wastewater treatment processes--identification of potential viral indicators. Sci. Total Environ. 2014, 488-489, 290–296.

(21) Symonds, E. M.; Nguyen, K. H.; Harwood, V. J.; Breitbart, M. Pepper mild mottle virus: A plant pathogen with a greater purpose in (waste)water treatment development and public health management. Water Res. 2018, 144, 1–12.

(22) Leibowitz, J.; Kaufman, G.; Liu, P. Coronaviruses: propagation, quantification, storage, and construction of recombinant mouse hepatitis virus. Curr Protoc Microbiol 2011, Chapter 15, Unit 15E.1.

(23) Lu, X.; Wang, L.; Sakthivel, S. K.; Whitaker, B.; Murray, J.; Kamili, S.; Lynch, B.; Malapati, L.; Burke, S. A.; Harcourt, J.; et al. US CDC Real-Time Reverse Transcription PCR Panel for Detection of Severe Acute Respiratory Syndrome Coronavirus 2. Emerging Infect. Dis. 2020, 26.

(24) Besselsen, D. G.; Wagner, A. M.; Loganbill, J. K. Detection of rodent coronaviruses by use of fluorogenic reverse transcriptase-polymerase chain reaction analysis. Comp Med 2002, 52, 111– 116.

(25) Haramoto, E.; Kitajima, M.; Kishida, N.; Konno, Y.; Katayama, H.; Asami, M.; Akiba, M. Occurrence of pepper mild mottle virus in drinking water sources in Japan. Appl. Environ. Microbiol. 2013, 79, 7413–7418.

(26) Zhang, T.; Breitbart, M.; Lee, W. H.; Run, J.-Q.; Wei, C. L.; Soh, S. W. L.; Hibberd, M. L.; Liu, E. T.; Rohwer, F.; Ruan, Y. RNA viral community in human feces: prevalence of plant pathogenic viruses. PLoS Biol. 2006, 4, e3.

(27) Gendron, L.; Verreault, D.; Veillette, M.; Moineau, S.; Duchaine, C. Evaluation of filters for the sampling and quantification of RNA phage aerosols. Aerosol Science and Technology 2010, 44, 893–901.

(28) Merkes, C. M.; Klymus K.E.; Allison M.J.; Goldberg C.; Helbing C.C.; Hunter M.E.; Jackson C.A.; Lance R.F.; Mangan A.M.; Monroe E.M.; Piaggio A.J.; Stokdyk J.P.; Wilson C.C.; Richter C. (2019) Generic qPCR Limit of Detection (LOD) / Limit of Quantification (LOQ) calculator. R Script. Available at: https://github.com/cmerkes/qPCR_LOD_Calc. DOI: https://doi.org/10.5066/P9GT00GB.

(29) Ståhlberg, A.; Kubista, M. The workflow of single-cell expression profiling using quantitative real-time PCR. Expert Rev Mol Diagn 2014, 14, 323–331.

(30) Benefield, A. E.; Skrip, L. A.; Clement, A.; Althouse, R. A.; Chang, S.; Althouse, B. M. SARS-CoV-2 viral load peaks prior to symptom onset: a systematic review and individual-pooled analysis of coronavirus viral load from 66 studies. medRxiv 2020.09.28.20202028; doi: https://doi.org/10.1101/2020.09.28.20202028.

(31) Linton, N. M.; Kobayashi, T.; Yang, Y.; Hayashi, K.; Akhmetzhanov, A. R.; Jung, S.-M.; Yuan, B.; Kinoshita, R.; Nishiura, H. Incubation Period and Other Epidemiological Characteristics of 2019 Novel Coronavirus Infections with Right Truncation: A Statistical Analysis of Publicly Available Case Data. J Clin Med 2020, 9.

(32) Liu, P.; Cai, J.; Jia, R.; Xia, S.; Wang, X.; Cao, L.; Zeng, M.; Xu, J. Dynamic surveillance of SARS-CoV-2 shedding and neutralizing antibody in children with COVID-19. Emerg. Microbes Infect. 2020, 9, 1254–1258.

(33) Han, M. S.; Seong, M.-W.; Heo, E. Y.; Park, J. H.; Kim, N.; Shin, S.; Cho, S. I.; Park, S. S.; Choi, E. H. Sequential analysis of viral load in a neonate and her mother infected with severe acute respiratory syndrome coronavirus 2. Clin. Infect. Dis. 2020, 71, 2236–2239.

(34) Bi, Q.; Wu, Y.; Mei, S.; Ye, C.; Zou, X.; Zhang, Z.; Liu, X.; Wei, L.; Truelove, S. A.; Zhang, T.; et al. Epidemiology and transmission of COVID-19 in 391 cases and 1286 of their close contacts in Shenzhen, China: a retrospective cohort study. Lancet Infect. Dis. 2020, 20, 911–919.

(35) R Core Team. R: A language and environment for statistical computing. R foundation for statistical computing, Vienna, Austria 2016. https://www.R-project.org/

(36) Fu, A.; Narasimhan, B.; Boyd, S. cvxr?: an r package for disciplined convex optimization. J Stat Softw 2020, 94.

(37) Ruckdeschel, P.; Kohl, M. General Purpose Convolution Algorithm inS 4 Classes by Means of FFT. J Stat Softw 2014, 59, 1–25.

(38) Gerrity, D.; Papp, K.; Stoker, M.; Sims, A.; Frehner, W. Early-pandemic wastewater surveillance of SARS-CoV-2 in Southern Nevada: Methodology, occurrence, and incidence/prevalence considerations. Water Research X 2021, 10, 100086.

(39) Chik, A. H. S.; Glier, M. B.; Servos, M.; Mangat, C. S.; Pang, X.-L.; Qiu, Y.; D’Aoust, P. M.; Burnet, J.- B.; Delatolla, R.; Dorner, S.; et al. Comparison of approaches to quantify SARS-CoV-2 in wastewater using RT-qPCR: Results and implications from a collaborative inter-laboratory study in Canada. J. Environ. Sci. (China) 2021, 107, 218–229.

(40) Pecson, B. M.; Darby, E.; Haas, C. N.; Amha, Y. M.; Bartolo, M.; Danielson, R.; Dearborn, Y.; Di Giovanni, G.; Ferguson, C.; Fevig, S.; et al. Reproducibility and sensitivity of 36 methods to quantify the SARS-CoV-2 genetic signal in raw wastewater: findings from an interlaboratory methods evaluation in the U.S. Environ. Sci.: Water Res. Technol. 2021.

(41) Ye, Y.; Ellenberg, R. M.; Graham, K. E.; Wigginton, K. R. Survivability, partitioning, and recovery of enveloped viruses in untreated municipal wastewater. Environ. Sci. Technol. 2016, 50, 5077–5085.

(42) La Rosa, G.; Iaconelli, M.; Mancini, P.; Bonanno Ferraro, G.; Veneri, C.; Bonadonna, L.; Lucentini, L.; Suffredini, E. First detection of SARS-CoV-2 in untreated wastewaters in Italy. Sci. Total Environ. 2020, 736, 139652.

(43) Ahmed, W.; Tscharke, B.; Bertsch, P. M.; Bibby, K.; Bivins, A.; Choi, P.; Clarke, L.; Dwyer, J.; Edson, J.; Nguyen, T. M. H.; et al. SARS-CoV-2 RNA monitoring in wastewater as a potential early warning system for COVID-19 transmission in the community: A temporal case study. Sci. Total Environ. 2021, 761, 144216.

(44) D’Aoust, P. M.; Mercier, E.; Montpetit, D.; Jia, J.-J.; Alexandrov, I.; Neault, N.; Baig, A. T.; Mayne, J.; Zhang, X.; Alain, T.; et al. Quantitative analysis of SARS-CoV-2 RNA from wastewater solids in communities with low COVID-19 incidence and prevalence. Water Res. 2021, 188, 116560.

(45) Gonzalez, R.; Curtis, K.; Bivins, A.; Bibby, K.; Weir, M. H.; Yetka, K.; Thompson, H.; Keeling, D.; Mitchell, J.; Gonzalez, D. COVID-19 surveillance in Southeastern Virginia using wastewater-based epidemiology. Water Res. 2020, 186, 116296.

(46) Wu, F.; Zhang, J.; Xiao, A.; Gu, X.; Lee, W. L.; Armas, F.; Kauffman, K.; Hanage, W.; Matus, M.; Ghaeli, N.; et al. SARS-CoV-2 Titers in Wastewater Are Higher than Expected from Clinically Confirmed Cases. mSystems 2020, 5.

(47) Huisman, J. S.; Scire, J.; Angst, D. C.; Neher, R. A.; Bonhoeffer, S.; Stadler, T. Estimation and worldwide monitoring of the effective reproductive number of SARS-CoV-2. medRxiv 2020.11.26.20239368; doi: https://doi.org/10.1101/2020.11.26.20239368.

