## Supporting Information for "Wastewater monitoring outperforms case numbers as a tool to track COVID-19 incidence dynamics when test positivity rates are high"

**Contents**

Text S1. Deconvolution to estimate daily number of infections

Table S1. RT-qPCR primers, probes and reaction conditions used in this work

Table S2. RT-qPCR assay efficiencies, limits of detection, and $R^2$ of standard curves

Table S3. Check-list of experimental details as requested by MIQE guidelines

Figure S1. Positivity rate in Switzerland during the first wave of the pandemic

Figure S2. Cq values measured in RNA extracts spiked with synthetic SARS-CoV-2 RNA reference material

Figure S3. SARS-CoV-2 N1 concentrations measured by RT-qPCR in biological and technical replicates

Figure S4. Recovery of three surrogate viruses simultaneously spiked into seven wastewater samples

Figure S5. Recovery of MHV (L ausanne and Lugano) or Sendai virus (Zurich) in longitudinal samples from the three WWTPs studied.

Figure S6. Daily load of PMMoV in Lausanne, Lugano and Zurich

Figure S7. SARS-CoV-2 N1 and N2 concentrations at each WWTP from February 26 to April 30, 2020

Figure S8. Comparison of the SARS-CoV-2 N1 and PMMoV RNA concentrations measured per ml of wastewater

References

**Text S1. Deconvolution to estimate daily number of infections**

The SARS-CoV-2 loads in wastewater and the daily confirmed cases are aggregated quantities. An aggregated quantity $A_t$ observed on day $t$ is the weighted sum all previous numbers of infections $I$. The weights $w$ are given as function of the time since infection, $\tau$:

$$A_t \approx \sum_{\tau=0}^{\infty} w(\tau) \cdot I_{t-\tau} \approx \sum_{\tau=0}^{\tau_{max}} w(\tau) \cdot I_{t-\tau}$$

As the weight decays with a larger $\tau$ the sum can be truncated at $\tau_{max}$ with a small error.

Based on the relationship from above we can estimate the infections $\boldsymbol{I} = (I_0, I_{1,} \dots, I_T \ )$ in a time interval $[0, \dots, T]$ from the set of observed aggregated quantities $\{A_t,\ t \in T_{obs}\}$. Note that the observation time points of $A_t$ may be irregularly spaced in time. The loss function to minimize over $\boldsymbol{I}$ is

$$\min_{\boldsymbol{I}} \sum_{t \in T_{obs}} \left( A_t - \sum_{\tau=0}^{T_{max}} w(\tau) \cdot I_{t-\tau} \right)^2 + \gamma \sum_{\tau=1}^{T_{max}} (I_\tau - I_{\tau-1})^2$$

with the constraint that all $I_\tau \geq 0$. The second term penalizes large deviations between successive infections and also ensures a unique solution. The larger the penalty term $\gamma$ is chosen, the smoother change the resulting estimates over time.

This model can be seen as a non-negative ridge regression. We implemented it using the R-package CVXR for convex optimization.

**Table S1**. RT-qPCR primers, probes and PCR reaction conditions used in this work.

| Virus | Ref | Primer name | Sequence (5'→3') | Primer concentration | Thermocycling protocol | Gene | Amplicon size (position) | Accession number |
|---|---|---|---|---|---|---|---|---|
| **PMMoV** | *1, 2* | PMMV-FP1-rev<br>PMMV-RP1<br>PMMV-Probe1 | 5'-GAGTGGTTTGACCTTAACGTTTGA-3'<br>5'-TTGTCGGTTGCAATGCAAGT-3'<br>5'-FAM-CCTACCGAAGCAAATG-MGB-3 | 0.4 µM<br>0.4 µM<br>0.2 µM | RT at 55ºC for 1h, 10 minutes at 95ºC followed by 45 cycles of 95ºC for 15s and 60ºC for 60s | Replication-associated protein | 68 (1878-1945) | NC_003630 |
| **Sendai virus** | This study | Sendai_F_2020<br>Sendai_R_ 2020<br>Sendai_P_2020 | 5'-GAGGTATAGGAGTCCCTGAAGT-3'<br>5'-GCTCTAGTCTGATGTCCCAAAG-3'<br>5'-FAM-TCCCGAGGCAGATAATGCACTGTT-ZEN/Iowa Black-3 | 0.4 µM<br>0.4 µM<br>0.2 µM | RT at 48ºC for 1h, 5 minutes at 95ºC followed by 45 cycles of 95ºC for 15s and 60ºC for 30s | L gene - RNA polymerase protein | 113 (13609-13726) | M30204 |
| **SARS-CoV-2 (N1)** | *3* | 2019-nCoV_N1-F<br>2019-nCoV_N1-R<br>2019-nCoV_N1-P | 5'-GACCCCAAAATCAGCGAAAT-3'<br>5'-TCTGGTTACTGCCAGTTGAATCTG-3'<br>5'-FAM-ACCCCGCATTACGTTTGGTGGACC-ZEN/Iowa Black-3' | 0.5 µM<br>0.5 µM<br>0.125 µM | RT at 55ºC for 1h, 10 minutes at 95ºC followed by 45 cycles of 95ºC for 15s and 55ºC for 30s | Nucleocapsid phosphoprotein | 72 (28287-28358) | NC_045512 |
| **SARS-CoV-2 (N2)** | *3* | 2019-nCoV_N2-F<br>2019-nCoV_N2-R<br>2019-nCoV_N2-P | 5'-TTACAAACATTGGCCGCAAA-3'<br>5'-GCGCGACATTCCGAAGAA-3'<br>5'-FAM-ACAATTTGCCCCCAGCGCTTCAG-ZEN/Iowa Black-3 | 0.5 µM<br>0.5 µM<br>0.125 µM | RT at 55ºC for 1h, 10 minutes at 95ºC followed by 45 cycles of 95ºC for 15s and 55ºC for 30s | Nucleocapsid phosphoprotein | 67 (29164-29230) | NC_045512 |
| **MHV** | *4* | MHV_F<br>MHV_R<br>MHV_P | 5'-GGAACTTCTCGTTGGGCATTATACT-3'<br>5'-ACCACAAGATTATCATTTTCACAACATA-3'<br>5'-FAM-ACATGCTACGGCTCGTGTAACCGAACTGT-BHQ-3' | 0.3 µM<br>0.3 µM<br>0.4 µM | RT at 50ºC for 1h, 5 minutes at 95ºC followed by 45 cycles of 95ºC for 15s and 60ºC for 60s | Membrane protein | 108 (29045-29152) | AY700211 |
| **ϕ6** | Adapted from *5* | Phi6_F<br>Phi6_R<br>Phi6_P | 5'-TGGCGGCGGTCAAGAG-3'<br>5'-GGATGATTCTCCAGAAGCTGCT-3'<br>5'-FAM-GTCGCAGGTCTGACACT-BHQ-3' | 0.4 µM<br>0.4 µM<br>0.08 µM | RT at 50ºC for 1h, 10 minutes at 95ºC followed by 40 cycles of 95ºC for 15s and 60ºC for 1 min | Nucleocapsid protein (Segment S) | 100 (429-528) | DQ785287 |

**Table S2**. RT-qPCR assay efficiencies, limits of detection, and $R^2$ of standard curves.

| Target | Efficiency [%] | LOD [gc/ml, (gc/reaction)] | Slope / intercept of standard curve | $R^2$ of pooled standard curve | Number of standard curves in pool |
|---|---|---|---|---|---|
| **SARS-CoV-2 N1** | 112 | 4.2 (13) | -3.07 / 38.24 | 0.97 | 21 |
| **SARS-CoV-2 N2** | 106 | 2.6 (8) | -3.18 / 39.14 | 0.95 | 16 |
| **MHV** | 98 | 3.2 (10) | -3.36 / 41.08 | 0.99 | 11 |
| **ϕ6** | 98 | 16 (50) | -3.37 / 39.52 | 0.99 | 3 |
| **Sendai virus** | 94 | 1.6 (5) | -3.47 / 38.87 | 0.99 | 4 |

**Table S3**. Check-list of experimental details as requested by MIQE guidelines[6]

| ITEM TO CHECK | Provided (Y/N) | CHECKLIST |
|---|---|---|
| **EXPERIMENTAL DESIGN** | | |
| Definition of experimental and control groups | Y | provided in methods section |
| Number within each group | Y | provided in methods section |
| **SAMPLE** | | |
| Description | Y | provided in methods section |
| Microdissection or macrodissection | N | N/A |
| Processing procedure | Y | provided in methods section |
| If frozen - how and how quickly? | Y | provided in methods section |
| If fixed - with what, how quickly? | N | N/A |
| Sample storage conditions and duration | Y | provided in methods section |
| **NUCLEIC ACID EXTRACTION** | | |
| Procedure and/or instrumentation | Y | provided in methods section |
| Name of kit and details of any modifications | Y | provided in methods section |
| Details of DNase or RNAse treatment | N | N/A |
| Contamination assessment (DNA or RNA) | Y | provided in methods section |
| Nucleic acid quantification | Y | provided in methods section and SI |
| Instrument and method | Y | provided in methods section |
| RNA integrity method/instrument | N | not done |
| RIN/RQI or Cq of 3' and 5' transcripts | N | not done |
| Inhibition testing (Cq dilutions, spike or other) | Y | provided in methods section |
| **REVERSE TRANSCRIPTION** | | |
| Complete reaction conditions | Y | provided in methods section |
| Amount of RNA and reaction volume | Y | provided in methods section |
| Priming oligonucleotide (if using GSP) and concentration | Y | provided in SI |
| Reverse transcriptase and concentration | Y | provided in SI |
| Temperature and time | Y | provided in SI |
| **qPCR TARGET INFORMATION** | | |
| Sequence accession number | Y | Published assays or provided in SI |
| Amplicon length | Y | Published assays or provided in SI |
| *In silico* specificity screen (BLAST, etc) | N | not provided |
| Location of each primer by exon or intron (if applicable) | N | N/A |
| What splice variants are targeted? | N | N/A |
| **qPCR OLIGONUCLEOTIDES** | | |
| Primer sequences | Y | provided in SI |
| Probe sequences | Y | provided in SI |
| Location and identity of any modifications | N | N/A |
| **qPCR PROTOCOL** | | |
| Complete reaction conditions | Y | provided in SI |
| Reaction volume and amount of RNA | Y | provided in methods section |
| Primer, (probe), Mg++ and dNTP concentrations | Y | provided in SI and according to kit instructions |
| Polymerase identity and concentration | Y | According to kit instructions |
| Buffer/kit identity and manufacturer | Y | provided in methods section |
| Additives (SYBR Green I, DMSO, etc.) | Y | provided in methods section |
| Complete thermocycling parameters | Y | provided in SI |
| Manufacturer of qPCR instrument | Y | provided in SI |
| **qPCR VALIDATION** | | |
| Specificity (gel, sequence, melt, or digest) | N | N/A |
| For SYBR Green I, Cq of the NTC | N | N/A |
| Standard curves with slope and y-intercept | Y | provided in SI |
| PCR efficiency calculated from slope | Y | provided in results section and SI |
| $R^2$ of standard curve | Y | provided in results section and SI |
| Linear dynamic range | N | Not assessed |
| Cq variation at lower limit | N | Not determined |
| Evidence for limit of detection | Y | provided in methods section |
| **DATA ANALYSIS** | | |
| qPCR analysis program (source, version) | Y | provided in methods section |
| Cq method determination | Y | provided in methods section |
| Outlier identification and disposition | N | N/A |
| Results of NTCs | Y | provided in results section |
| Justification of number and choice of reference genes | N | N/A |
| Description of normalisation method | N | N/A |
| Number and concordance of biological replicates | Y | provided in methods section |
| Number and stage (RT or qPCR) of technical replicates | Y | provided in methods section |
| Repeatability (intra-assay variation) | Y | provided in results section and SI |
| Statistical methods for result significance | Y | provided in methods section |
| Software (source, version) | Y | provided in methods section |

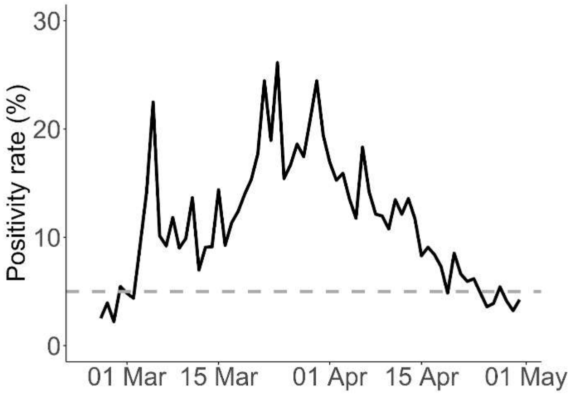


**Figure S1**: Positivity rate in Switzerland during the first wave of the pandemic. Data source: Swiss Federal Office of Public Health.

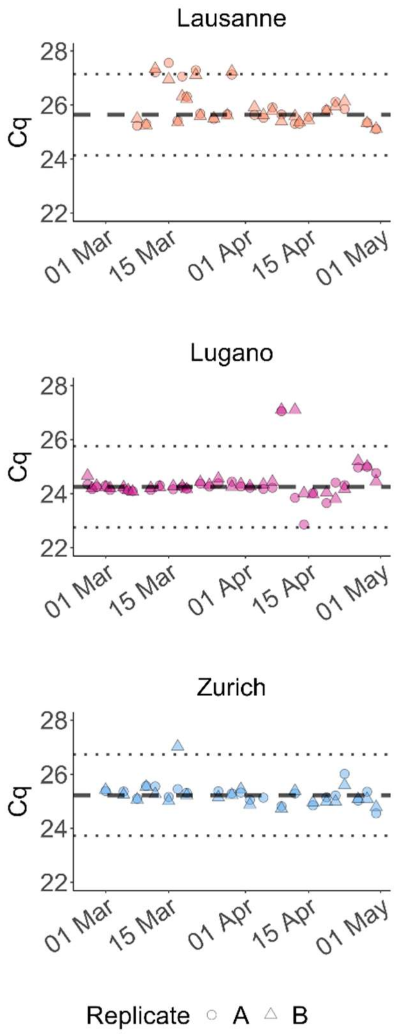


**Figure S2**. Cq values measured in RNA extracts spiked with synthetic SARS-CoV-2 RNA reference material. Dashed lines indicate the median Cq for a given site. Dotted lines indicate the median ± 1.5 Cq, considered as the range with minimal PCR inhibition.

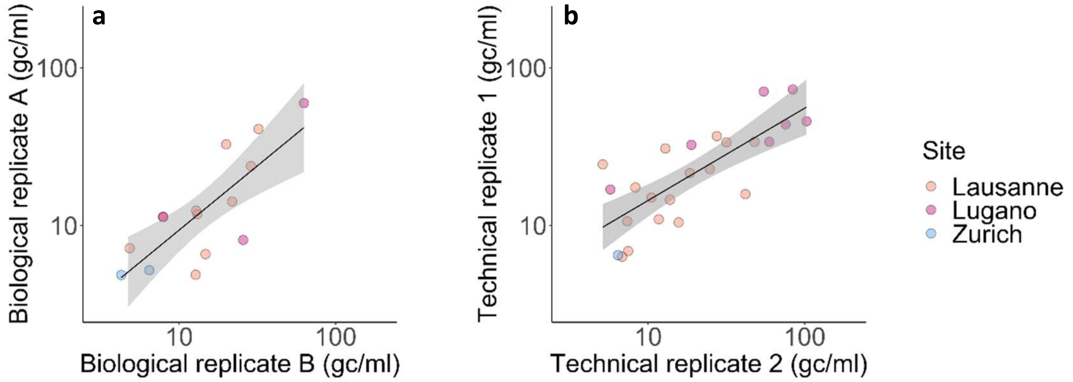


**Figure S3**. SARS-CoV-2 N1 concentrations measured by RT-qPCR (expressed as gc/reaction) in biological (a) and technical (b) replicates. One PCR reaction corresponds to 5 µl of RNA extract, or 3.125 ml of wastewater. The shaded areas indicate the standard error associated with the regression line. Only measurements > LOD were considered.

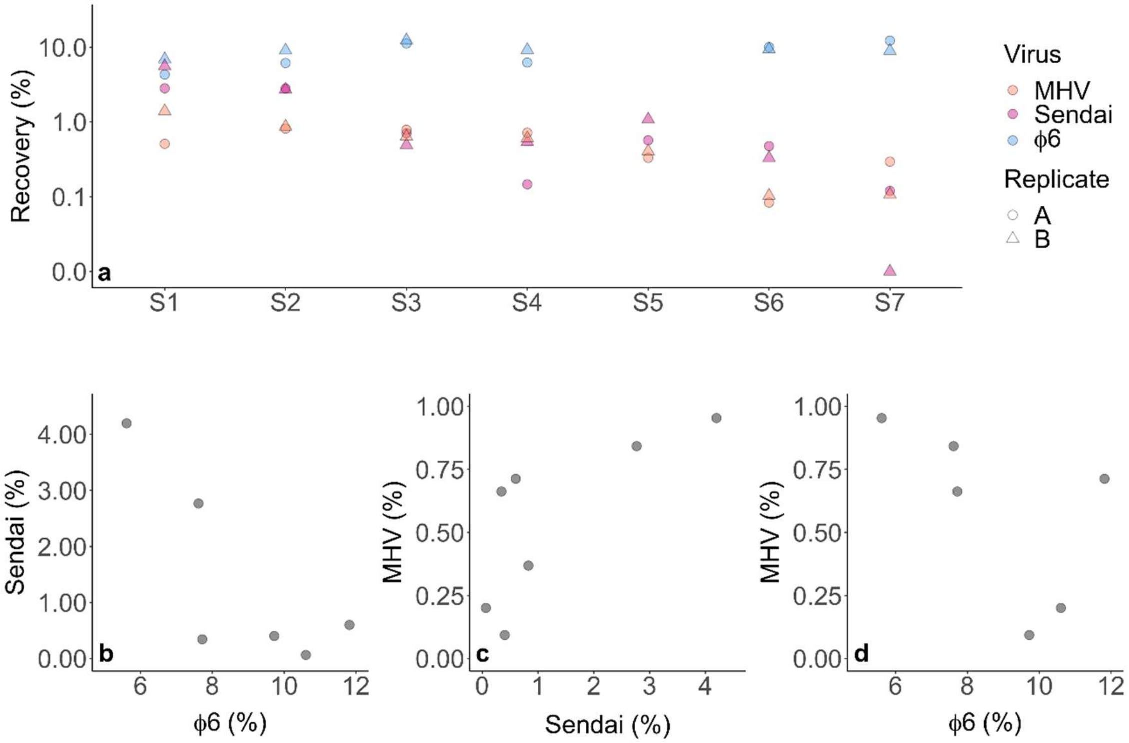


**Figure S4**: Recovery of three surrogate viruses (MHV, Sendai virus and Φ6) simultaneously spiked into seven wastewater samples. Each wastewater sample was processed and analyzed in duplicate. Panel a) Overview over all recoveries measured. Panels b-d) Paired comparison between recoveries measured by the three surrogates considered.

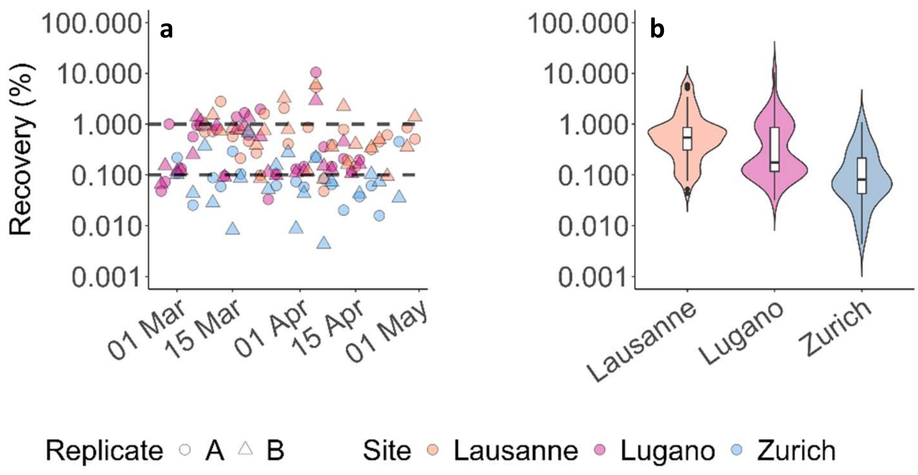


**Figure S5**. a) Recovery of MHV (Lausanne and Lugano) or Sendai virus (Zurich) in longitudinal samples from the three WWTPs studied. Each sample was processed and analyzed in duplicate. b) Violin plots show the kernel probability density (i.e. the proportion of data located along the range of recovery values, smoothed by a kernel estimator). Inside, boxplots show the median and interquartile range.

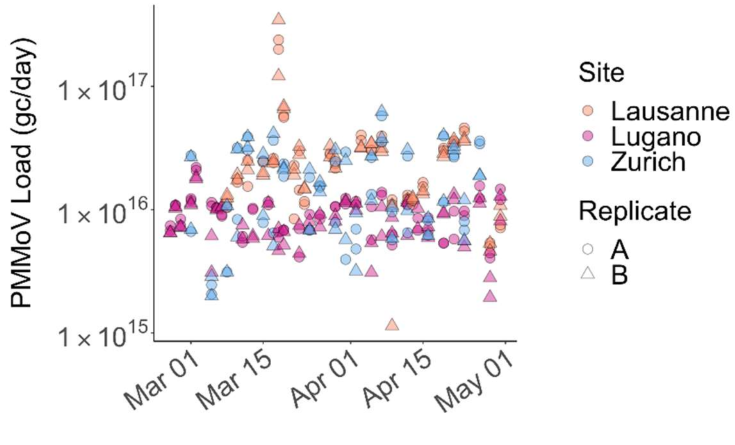


**Figure S6.** Daily load of PMMoV in Lausanne, Lugano and Zurich. Each sample was processed and analyzed in duplicate.

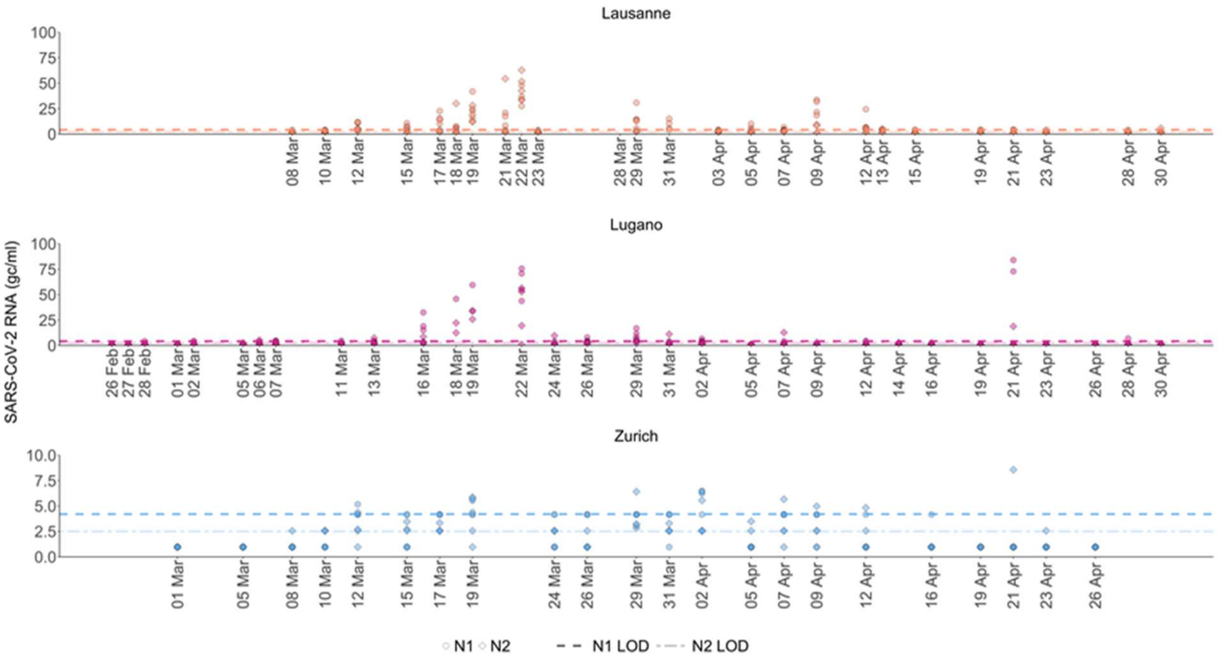


**Figure S7**. SARS-CoV-2 N1 and N2 concentrations at each WWTP from February 26 to April 30, 2020. All biological and technical replicates are plotted as individual data points. Dashed lines indicate the LOD for the N1 (4.2 gc/ml) and N2 (2.6 gc/ml) in wastewater.

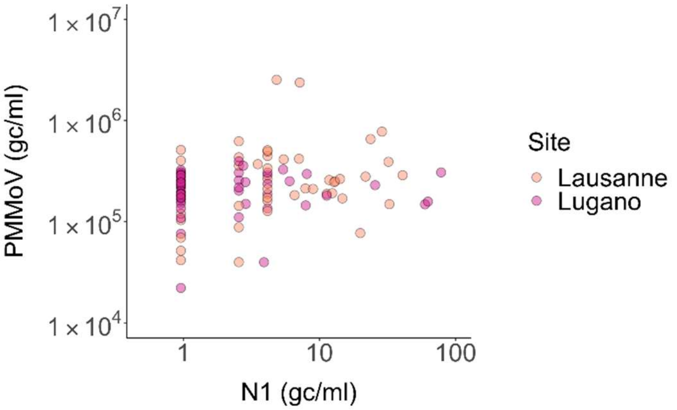


**Figure S8**. Comparison of the SARS-CoV-2 N1 and PMMoV RNA concentrations measured per ml of wastewater. Because the Zurich WWTP mostly yielded N1 concentrations < LOD, this site was not included in the graph.

**References**

(1) Haramoto, E.; Kitajima, M.; Kishida, N.; Konno, Y.; Katayama, H.; Asami, M.; Akiba, M. Occurrence of pepper mild mottle virus in drinking water sources in Japan. *Appl. Environ. Microbiol.* **2013**, *79*, 7413–7418.

(2) Zhang, T.; Breitbart, M.; Lee, W. H.; Run, J.-Q.; Wei, C. L.; Soh, S. W. L.; Hibberd, M. L.; Liu, E. T.; Rohwer, F.; Ruan, Y. RNA viral community in human feces: prevalence of plant pathogenic viruses. *PLoS Biol.* **2006**, *4*, e3.

(3) Lu, X.; Wang, L.; Sakthivel, S. K.; Whitaker, B.; Murray, J.; Kamili, S.; Lynch, B.; Malapati, L.; Burke, S. A.; Harcourt, J.; et al. US CDC Real-Time Reverse Transcription PCR Panel for Detection of Severe Acute Respiratory Syndrome Coronavirus 2. *Emerging Infect. Dis.* **2020**, *26*.

(4) Besselsen, D. G.; Wagner, A. M.; Loganbill, J. K. Detection of rodent coronaviruses by use of fluorogenic reverse transcriptase-polymerase chain reaction analysis. *Comp Med* **2002**, *52*, 111–116.

(5) Gendron, L.; Verreault, D.; Veillette, M.; Moineau, S.; Duchaine, C. Evaluation of filters for the sampling and quantification of RNA phage aerosols. *Aerosol Sci. Technol.* **2010**, *44*, 893–901.

(6) Bustin, S. A.; Benes, V.; Garson, J. A.; Hellemans, J.; Huggett, J.; Kubista, M.; Mueller, R.; Nolan, T.; Pfaffl, M. W.; Shipley, G. L.; et al. The MIQE guidelines: minimum information for publication of quantitative real-time PCR experiments. *Clin. Chem.* **2009**, *55*, 611–622.
